# The impact of measures set by national regulatory authority to enhance affordability of medicines in Sudan: when good intention leads to worse outcomes

**DOI:** 10.1101/2024.03.13.24304248

**Authors:** Gamal Khalafalla Mohamed Ali, Yasir Abubaker Mohamed Abuelrish, Abubakr Abdelraouf Alfadl, Mohamed Abdelrahman Mohamed Shigidi

## Abstract

**Introduction:** The aim of this paper is to assess the impact of the measures and procedures imposed by the National Medicines and Poisons Board (NMPB) on the availability of safe, effective and quality medicines of affordable price on the Sudanese market.

**Methods:** This is a descriptive study in which pharmacists, who were responsible for the regulatory affairs within their importing and locally manufacturing medicine companies, were asked to complete a 45-question online survey using the Google application, covering relevance and challenges of medicines quality and pricing system. A link to the data collection tool website was sent to all members of the Sudanese Society of Regulatory Affairs Pharmacists through WhatsApp. The survey was available on 6th May 2020 through 6th June 2020. Descriptive statistics were used to report results.

**Results:** Survey respondents were 70 regulatory affairs’ pharmacists. 38% of participants believe that the technical procedures adopted by the NMPB do not achieve the goal of establishing Medicine Regulatory Authorities as described by World Health Organization. Almost all respondents believe that Sudan current economic situation, including the scarcity of foreign currency, has greatly impacted the availability of quality-assured medicines in pharmacies. Participants said that the situation is exacerbated by the intervention of NMPB in determining the exchange rate and controlling the medicine prices.

**Conclusion:** The NMPB should consider options for balancing patient access to quality medicines, and reasonable pricing policies that encourage the local pharmaceutical manufacturing to flourish and a steady flow of quality-assured medicines from abroad to the Sudan market.

## Introduction

World Health Organization (WHO) reported that ‘Shortages of essential drugs are becoming increasingly frequent globally, burdening health systems with additional costs and posing risks to the health of patients who fail to receive the medicines they need’ [p.180]^(1)^. The risk for patients results of non-treatment, under-treatment, possible medication errors from attempt to substitute non-available medicines and low-quality medicines^(1)^. The medicine shortages are also affecting prescribers and dispenser ^(2–5)^.

The role of National Medicine Regulatory Authorities (NMRAs) that includes, but not limited to, registration of medicines and licensing of pharmaceutical establishments has been identified by WHO^(6)^. In Sudan, National Medicines and Poisons Board (NMPB), the NMRA, also controls medicine prices. The impact of measures applied by NMRAs to assure safety, efficacy and quality on access to medicines is reported elsewhere (see, for example,^(1, 7, 8)^). However, medicine shortages as a result of price control, to our knowledge, are not well researched, especially in low- and middle-income countries.

After the separation of South Sudan in July 2011, Sudan lost 75% of oil resources^(9)^. This secession affected the availability of foreign currency in Sudan. The devaluation of the Sudanese pound (SDG) during 2012 jumped to 57% and shot up to more than 410% by the end of 2020. The black market rate steadily increases. As a result, the pharmaceutical companies experiencing barriers to access the foreign currency for importing medicines and their raw materials at the official exchange rate.

**Figure I:**
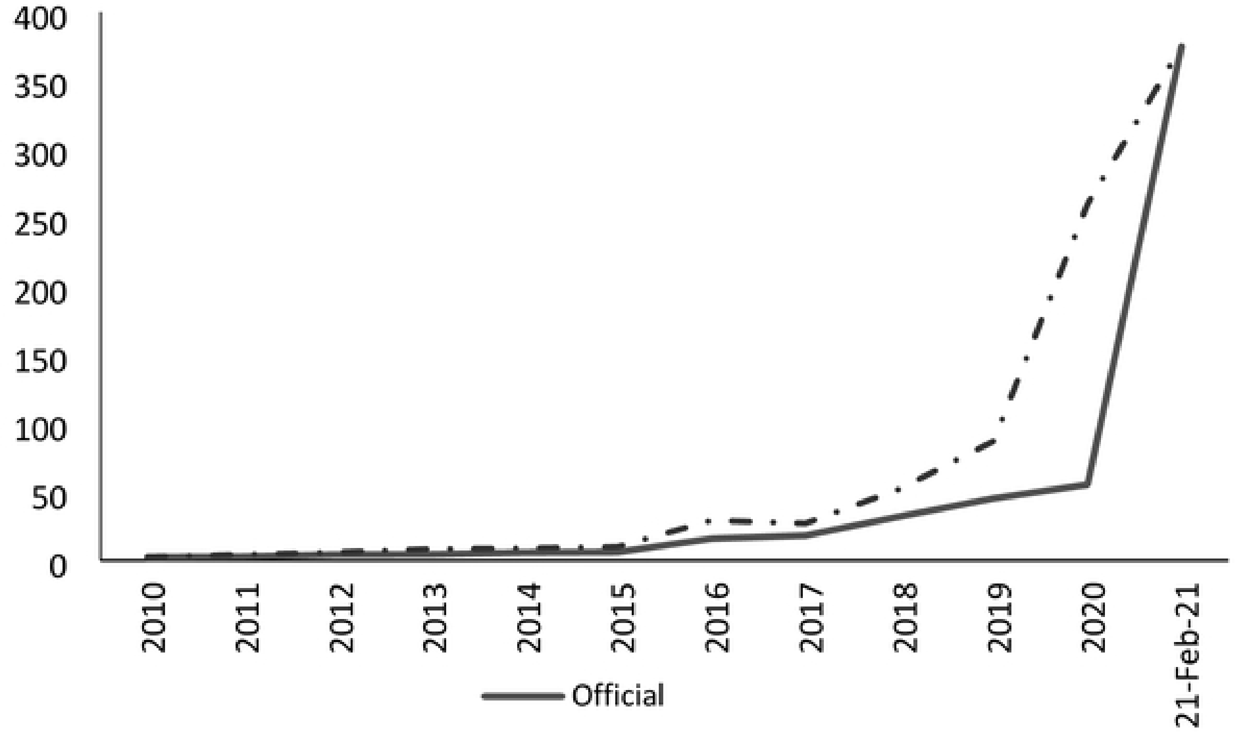
Exchange Rate of US$ to SDG: Comparison between Official and Black Market.

Since early 2013, the Sudanese media, especially newspapers, regularly claim that people are suffering from great shortages in life-saving medicines in the private markets. The newspapers and activists on social media reported patients and their relatives are wasting their time and money going from one pharmacy to another in search for their prescribed medicines. The lead column writers appealed for the government to intervene.

Against this situation and in response to the officials’ directives and the media to increase the number of registered medicines, NMPB has adopted a number of measures to control the impact of devaluation of local currency on the prices of medicines. These measures are: the NMPB has replaced the pre-marketing quality control laboratory testing by collecting samples for quality tests upon the arrival of the first consignment of the new registered medicines. The NMPB also has abolished the compulsory submission of the medicine dossier in the format of Common Technical Document (CTD) that has been used by the members of the International Council for Harmonization. Additionally, the exclusive agency rights were cancelled by the Presidential Decree No. 669 in October 28, 2018. Further, the NMBP exempted a number of countries that have no stringent NMRAs from inspection. Furthermore, the NMPB has revoked the ceiling resolution. According to this resolution the maximum number to be registered of a multi-source (generic) pharmaceutical product of the same dosage form and strength was ten. Moreover, the NMPB canceled the positive and negative lists, which included high priority medicines in registration because they were not available on the market, and the medicines of which sufficient numbers have been registered respectively. Furthermore, the NMPB obligated importing companies to reduce CIF (Cost, Insurance and Freight) prices for registered medicines. The NMPB also agreed with the national medicine manufacturers to charge same price to all generic medicines. Finally, by a Circular No. 4 dated 25^th^ July 2018, the NMPB has limited the quantities that pharmaceutical companies allowed to sell every month.

By easing the requirement for medicine registration, the NMPB announced the registration of 1,000 items in 2014 and about 900 items in 2018. Despite these measures and the registration of unprecedented number of generic medicines, the availability of quality-assured medicines at affordable prices was at stake. In fact, the situation was worsened and the access to medicines became a real challenge to the patients.

This paper aims to find out the impact of the NMPB’s measures on the availability of safe, effective and quality medicines of affordable price on the Sudanese market from the perspective of the members of the Sudanese Society of Regulatory Affairs Pharmacists (SSRAP). Therefore, it will contribute to the provision of reliable information that will help NMPB and the NMRAs in developing countries with similar socio-economic situations, especially in Sub-Saharan Africa, to formulate appropriate policies and make informed-decisions that retain patients’ access to life-saving medicines.

## Method

The SSRAP was first established in 2018. It constitutes all pharmacists who are responsible for regulatory affairs within their medicines importing or manufacturing companies. A questionnaire was designed to know underlying causes of medicines shortages, as viewed from a perspective of regulatory affairs pharmacists in Sudan. The questions were compiled, evaluated, reviewed and approved by all authors.

Participants were asked to complete a 45-question online survey using the Google application, covering relevance and challenges of medicines quality and pricing system. The questions were of various types, ranging from multiple choices to multiple responses and additional free text answers were possible. For example, in most of the questions, the participants were asked to indicate their level of agreement or disagreement using a 5-point Likert-scale (strongly agree = 1 point to strongly disagree = 5 points). The survey instrument was pilot tested on 4 of the SSRAP’s members before the questions were sent out. A WhatsApp message was sent to all members of the SSRAP with a brief preamble on the topic of “impact of the NMPB measures on the flow and availability of medicines in Sudan” and a link inviting them to participate in the survey. Apart from the gender, date of employment and the type of medicine company (whether a manufacturer or an importer), no personal identifying information was collected. The survey was opened on 6^th^ May 2020 and was closed on 6^th^ June 2020, with additional WhatsApp messages to pharmacists to encourage participation. Descriptive statistics were compiled for each question. The data were analyzed automatically using Microsoft Office Excel 2016.

## Results

### Demographic characteristics of participants

The registered members of the SSRAP at the time of the study were 100 pharmacists. In total, 70 pharmacists participated in the survey (response rates of 70%). As shown in Table 1, nearly 70% of the study participants disclosed their gender, just fewer than 60% of whom were females. The dates of participants’ employment in their current jobs spread throughout the past 28 years.

**Table 1:** Demographic characteristics of the respondents.

| **Characteristics** | **Response rate n (%)** |
| --- | --- |
| **<u>Gender</u>** |  |
| Male | 29 (41.4) |
| Female | 41 (58.6) |
| **<u>Job</u>** |  |
| Regulatory affairs | 29 (41.4) |
| Registration officer | 14 (20) |
| Pharmacist | 2 (3) |
| Manager (company) | 11 (15.7) |
| Manager (industry) | 6 (8.6) |
| General Manager | 5 (7.1) |
| Technical consultant | 1 (1.4) |
| Medical representative | 1 (1.4) |
| Missing | 1 (1.4) |
| **<u>Employer</u>** |  |
| Drug importing company | 51 (72.9) |
| Pharmaceutical manufacturer | 12 (17.1) |
| Drug importing and manufacturing company | 7 (10.0) |
| **<u>No. of years in the current job</u>** |  |
| < 5 | 27 (38.6) |
| 5 – 9 | 24 (34.3) |
| 10 – 14 | 10 (7.0) |
| 15 – 19 | 6 (8.6) |
| ≥ 20 | 3 (4.3) |

### Quality of medicines

Surprisingly, 38% of those who participated in this study believe that the technical procedures that the NMPB has adopted in the past 10 years do not achieve the goal of establishing NMRAs as described above. Nevertheless, about half of the participants reported that the NMPB’s actions ensure the safety, efficacy and quality of medicines circulated in Sudan. More than 40% of pharmacists who participated in the study thought that the NMPB’s measures do not prohibit the circulation of substandard and falsified (SF) medicines. However, just below 30% of participants reported their confidence in the NMPB’s measures in protecting public health against SF medicines, with 60% of participants reported that the high number of medicine wholesalers will undermine the regulatory system in Sudan and more likely will lead to circulation of SF medicines in the country legitimate supply chain of health products. Finally, 55% of the participants were dissatisfied with the performance of regulatory institutions (i.e. NMPB and its state branches) in Sudan. Details of the responses of the participants on various statements regarding their perspective on the impact of NMPB’s measures on the availability of quality medicines are shown in Table 2 below.

**Table 2:**
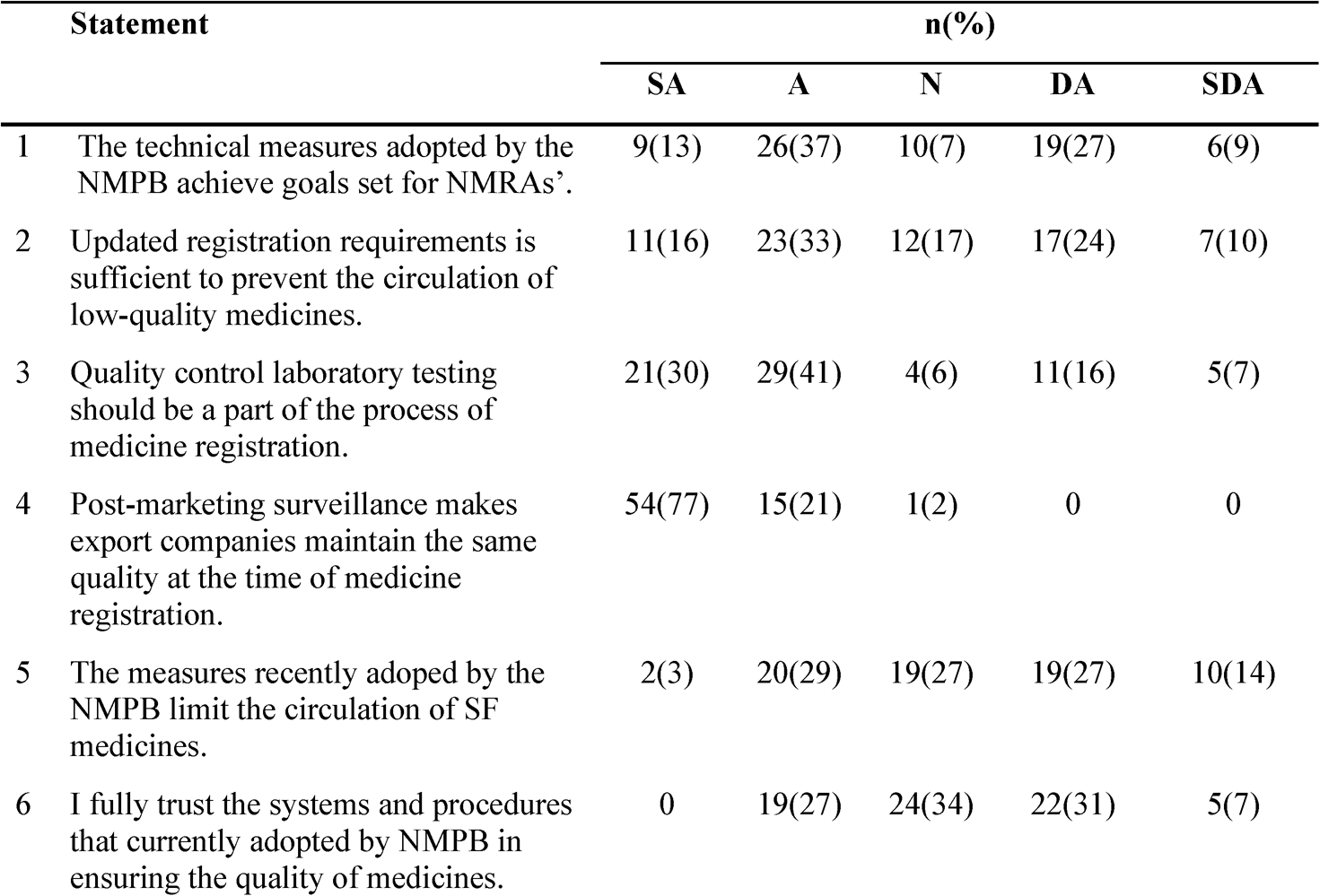
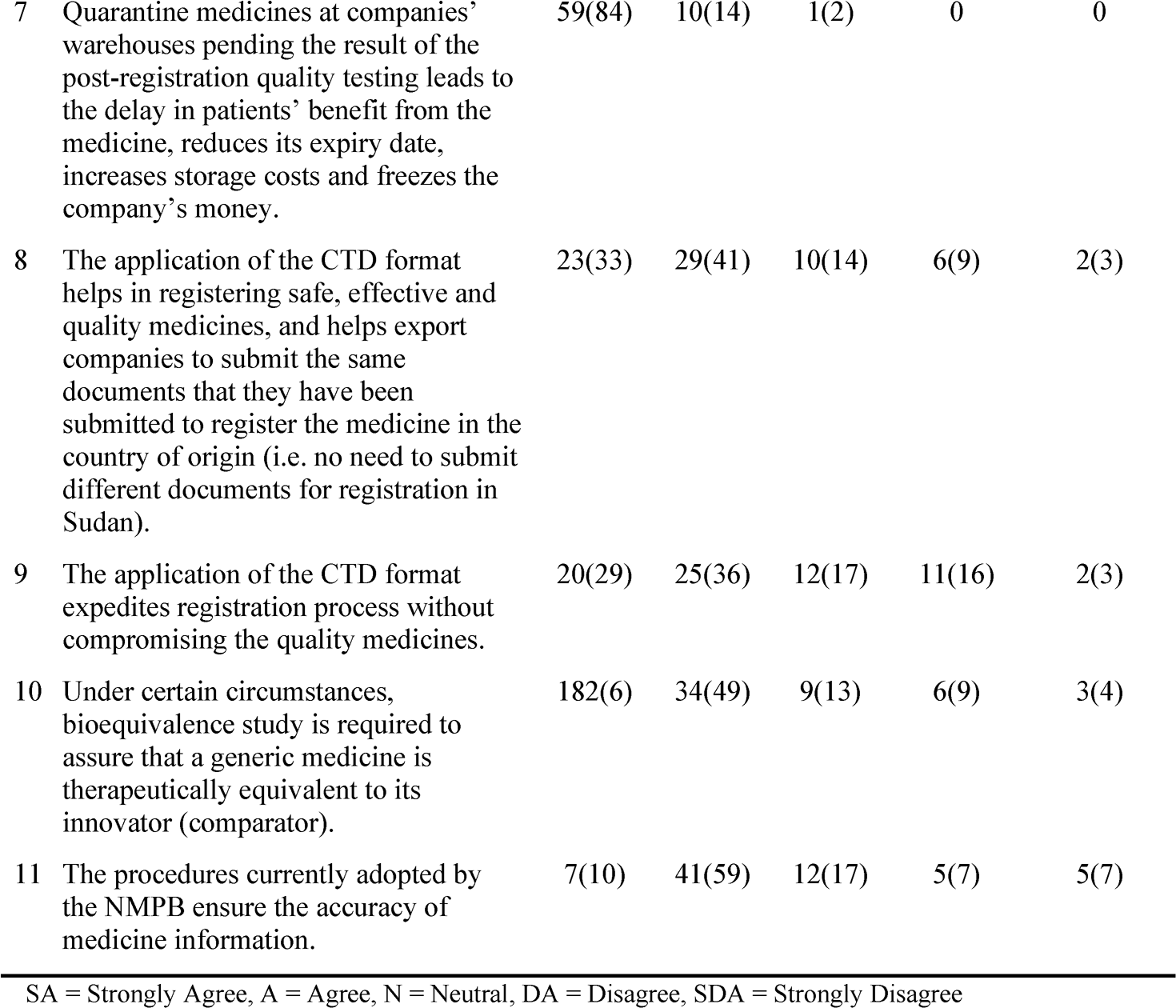
Participants’ perspective about the quality of medicines.

| Statement | n(%) |  |  |  |  |
| --- | --- | --- | --- | --- | --- |
|  | SA | A | N | DA | SDA |
| 1 The technical measures adopted by the NMPB achieve goals set for NMRAs'. | 9(13) | 26(37) | 10(7) | 19(27) | 6(9) |
| 2 Updated registration requirements is sufficient to prevent the circulation of low-quality medicines. | 11(16) | 23(33) | 12(17) | 17(24) | 7(10) |
| 3 Quality control laboratory testing should be a part of the process of medicine registration. | 21(30) | 29(41) | 4(6) | 11(16) | 5(7) |
| 4 Post-marketing surveillance makes export companies maintain the same quality at the time of medicine registration. | 54(77) | 15(21) | 1(2) | 0 | 0 |
| 5 The measures recently adopted by the NMPB limit the circulation of SF medicines. | 2(3) | 20(29) | 19(27) | 19(27) | 10(14) |
| 6 I fully trust the systems and procedures that currently adopted by NMPB in ensuring the quality of medicines. | 0 | 19(27) | 24(34) | 22(31) | 5(7) |

|  |  |  |  |  |  |  |
| --- | --- | --- | --- | --- | --- | --- |
| 7 | Quarantine medicines at companies' warehouses pending the result of the post-registration quality testing leads to the delay in patients' benefit from the medicine, reduces its expiry date, increases storage costs and freezes the company's money. | 59(84) | 10(14) | 1(2) | 0 | 0 |
| 8 | The application of the CTD format helps in registering safe, effective and quality medicines, and helps export companies to submit the same documents that they have been submitted to register the medicine in the country of origin (i.e. no need to submit different documents for registration in Sudan). | 23(33) | 29(41) | 10(14) | 6(9) | 2(3) |
| 9 | The application of the CTD format expedites registration process without compromising the quality medicines. | 20(29) | 25(36) | 12(17) | 11(16) | 2(3) |
| 10 | Under certain circumstances, bioequivalence study is required to assure that a generic medicine is therapeutically equivalent to its innovator (comparator). | 182(6) | 34(49) | 9(13) | 6(9) | 3(4) |
| 11 | The procedures currently adopted by the NMPB ensure the accuracy of medicine information. | 7(10) | 41(59) | 12(17) | 5(7) | 5(7) |

SA = Strongly Agree, A = Agree, N = Neutral, DA = Disagree, SDA = Strongly Disagree

Only 49% of respondents believe that the new registration requirements are sufficient to protect the Sudanese market against SF medicines. And as a result, almost 80% of the respondents believe that the quality of medicines in the community pharmacies is fairly appropriate, while 16% of them describe the quality of medicines in community pharmacies as low.

As expected, more than 70% of participants believe that samples for quality control laboratory testing should be available as a part of the medicine dossier submitted to the NMPB for premarket evaluation. On the other hand, almost all participants (99%) believe that the NMPB should continue quality control laboratory testing of samples selected randomly from pharmacies. Again almost all participants (99%) believe that the registration and importation of medicines without pre-quality testing, and then quarantine them at importing companies’ warehouses pending the analysis leads to delay in the access to the medicines, reduces the shelf life, increases the warehousing costs, tied-up the funds, and depletes the capital of the companies due to inflation of the local currency.

The study revealed that one-third of the participants had experience or aware of cases when the NMPB at some point in the past authorized importing companies to distribute their medicines without waiting for the results of the laboratory test due to unexpected laboratory delays, and later on upon completion of the laboratory test the samples were found not conforming to the specifications and the company was unable to recall the item in question.

Three quarters of the respondents believe that the application of the CTD, would result in the registration of safe, effective and quality-assured medicines and 65% agreed that the adoption of the CTD could expedite registration of medicines. The study showed that although for certain types of generic medicines, bioequivalence study is the only tool available to prove that these generics are bioequivalent to their originators, this study is not mandatory in the new registration requirements. When asked about this matter, 75% of the participants agreed that the bioequivalence studies should be mandatory for generic medicines whose safety and efficacy cannot be confirmed by the comparative dissolution test.

### Impact of the administrative measures

The participants were asked whether any number of license holders may be authorized to market the same active ingredient of the same dosage form and strength, or whether the number for each should be restricted. The majority (about 75%) believes that having limited number of license holders is important. The proponents (20% of participants) agreed on exemption of the innovator and locally manufactured medicines from this ceiling. According to the participants, the limited number of license holders have several advantages, such as encouraging importing and manufacturing companies to register medicines that are not existing on the market, limiting unethical promotion and marketing malpractices, increasing the company’s market share, and saving the limited NMPB’s resources and the time of its employees to review dossiers of medicines that are not registered in Sudan (Table 3).

**Table 3:** Participants’ perception on NMPB administrative measures.

|  |  | n(%) |  |  |  |  |
| --- | --- | --- | --- | --- | --- | --- |
|  |  | SA | A | N | DA | SDA |
| 1 | Pharmacies, not patients, benefit from the free medicines (bonus), which motivates mal-practices to maximize profits at the expense of patients' well-being. | 29(41) | 26(37) | 6(9) | 8(11) | 1(2) |
| 2 | Set a ceiling for the number of registered generic medicines that excludes innovator and locally produced medicines, will save resources and time for the registration of generic medicines that do not exist on Sudan market. | 28(40) | 23(33) | 3(4) | 13(19) | 3(4) |
| 3 | The approval of proforma invoice and NMPB's certificate of no objection are not important for the quality of the medicines. | 22(31) | 21(30) | 5(7) | 17(24) | 5(7) |
| 4 | Generally, the NMPB's administrative measures in the past few years, such as setting the exchange rate, do not achieve the goal for which MRAs are established. | 25(36) | 31(44) | 9(13) | 4(6) | 1(2) |
| 5 | The large number of distributors may lead to the circulation of SF medicines. | 18(26) | 24(34) | 15(21) | 10(14) | 3(4) |

SA = Strongly Agree, A = Agree, N = Neutral, DA = Disagree, SDA = Strongly Disagree

As presented in Table 3, overall, the majority (80%) of participants considers that the administrative processes (such as NMPB’s approval of the proforma invoice; NMPB’s certificate of no objection; and a request for clearance after the arrival of the medicines) enforced by the NMPB do not achieve the goal of establishing NMRA that aims to ensure the safety, efficacy and quality of medicine. Instead, these documents are obstacles to the flow of medicines to Sudan. The documents can only be replaced by the clearance permission to ensure that the medicines have marketing authorization, are imported from the registered source and with the same specifications and CIF prices.

### Medicine prices

Almost all respondents believe that the scarcity of foreign currency has greatly impacted the availability of quality-assured medicines in pharmacies. Participants said that the situation is exacerbated by the intervention of NMPB in determining the exchange rate. Since the Central Bank of Sudan moved the official exchange rate from SDG 30 to about SDG 47 for one US$ in November 2018 and from SDG 47 to about SDG 55 per one US$ at the beginning of 2020, the NMPB had until recently priced medicines at an exchange rate lower than the formal one announced by the Central Bank of Sudan. For example, the NMPB set the sale price for pharmacies at the exchange rate of SDG 30, at a time when the official exchange rate was SDG 47. After 4 months and in early 2019, the NMPB agreed to adopt the official exchange rate as announced by the Central Bank. However, the official exchange rate quickly moved to SDG 55 for one US$. Similarly, and for the second time, the NMPB refused to adopt the new rate for almost two months, before adopting it. The majority (89%) of the participants said that the NMPB’s insistence in an exchange rate below the official rate and the compelling of medicine companies to do so weakened the ability of these companies to continue importing and manufacturing medicines.

Details of the responses of the participants about their perspective on the impact of medicines pricing measures on the availability of affordable, quality medicines are shown in Table 4 below:

**Table 4:** Responses of the participants on medicine pricing.

|  | Statements | n(%) |  |  |  |  |
| --- | --- | --- | --- | --- | --- | --- |
|  |  | SA | A | N | DA | SDA |
| 1 | The exchange rate used by NMPB to set wholesale price in SDG has weakened companies' importation capacity. | 51(73) | 18(26) | 1(2) | 0 | 0 |
| 2 | The NMPB should involve stakeholders in preparation of regulations, pricing policies or adoption of the exchange rate. | 51(73) | 17(24) | 0 | 2(3) | 0 |
| 3 | The high CIF at which some companies registered their medicines, led to distortions in medicine prices, which reflected in several mal-practices (e.g. the bonus). | 21(30) | 31(44) | 16(23) | 2(3) | 0 |
| 4 | Before revoking it in 2017, the NMPB did not enforce the pricing regulations 2010, especially the article on the exchange rate. The consequence was a disruption in the medicine flow to Sudan. | 39(56) | 12(17) | 8(11) | 0 | 0 |
| 5 | If NMPB had applied enforced the medicine pricing regulations 2010, it would have solved the recent problem of medicine pricing. | 20(29) | 22(31) | 21(30) | 6(9) | 1(2) |
| 6 | Scarcity of foreign currency at official exchange rates decreases the availability of quality medicines at community pharmacies. | 69(98) | 1(2) | 0 | 0 | 0 |

SA = Strongly Agree, A = Agree, N = Neutral, DA = Disagree, SDA = Strongly Disagree

Additionally, the new method of calculating the CIF price of medicines has negatively affected the flow of medicines to Sudan (Table 5), where 27% of the respondents reported that their companies stopped importing 30 types of innovative medicines after the NMPB has reduced their approved CIF prices which have been registered long time ago. Similarly, more than one-fifth of the participants reported that their companies could not import newly registered 23 innovative medicines because the CIF prices given by the NMPB were too low. On the other hand, 47% and 43% of the participants indicated that their companies stopped importing 49 generic medicines to the market and 294 recently registered generics after the reduction of the already approved or newly given CIF prices respectively.

**Table 5:** Consequences of NMPB’s pricing policies: participants’ experiences.

| Questions | n(%) |  |
| --- | --- | --- |
|  | Yes | No |
| 1 Has your company stopped importation of a brand medicine "innovator" in objection to the CIF price approved by the NMPB? | 19(27) | 51(73) |
| 2 During the past ten years, has your company registered an innovator medicine and did not import it since the registration, because the CIF price set by the NMPB was acceptable? | 16(23) | 54(77) |
| 3 Has your company stopped importation of a generic medicine in objection to the CIF price set by the NMPB? | 33(47) | 37(53) |
| 4 During the past ten years, has your company registered a generic medicine and did not import it, in objection to the CIF price set by the NMPB? | 40(57) | 30(43) |
| 5 Since 2013, has the factory, where you work, stopped production of a medicine in objection to the price set by the NMPB? | 12(17) | 58(83) |
| 6 At some point, the NMPB gave the same price for the same locally manufactured medicines. Does this policy solve the problem of pricing the locally produced medicines? | 10(14) | 60(86) |
| 7 Do you aware of the regulations of pricing registered pharmaceuticals issued in 2010, and canceled by the NMPB few years ago? | 39(56) | 31(44) |
| 8 Do you think restating and enforcing of the regulations of medicine pricing 2010 will solve the long-lasting problem of medicine pricing? | 35(50) | 35(50) |

To keep pace with the NMPB’s measures, participants reported that their companies have taken a number of strategies to avoid losses including decreasing the deferred payment period; requiring cash payment; selling in very small quantities; downsizing workforce; reducing employees’ benefit packages; and reducing the quantities of free-of-charge medicines. The participants believe that these short-term companies’ strategies have negative effects on the availability of medicines in Sudan because foreign manufacturers do not offer their local agents deferred payment anymore, due to the delays in paying back the previous orders. In addition, the recently applied prepayment method causes losses to medicine importing companies and depletes their capital by inflation. Further, the importing companies struggle to make prepayments, due to the scarcity of foreign currencies at the official exchange rates. Finally, the foreign manufacturers do not include Sudan agents in their manufacturing plans, because they are unable to withdraw the previous allocated quantities.

Local pharmaceutical manufacturers were not affected much, with 17% of respondents explaining that local manufacturers had suspended the production of more than 50 items in protest against the new low prices set by the NMPB. All participants from local manufacturers object to the new method of pricing, under which a single price is set for the locally manufactured similar pharmaceutical products. The participants argued that different manufacturers have different production costs because the cost of raw materials varies among manufacturers; the quality and the sources of raw materials are not the same; the production capacities are different and accordingly the volume of the production varies; the quality of the packing materials varies among manufacturers; the packing method varies from factory to factory; the number of medicines produced is different; and fixed costs, such as employees’ benefit packages, vary among manufacturers.

The participants thought that the NMPB does not keep pace with the inflation and insist on pricing medicines at an exchange rate below the official exchange rate due to the following reasons:

a. The NMPB does not want to recognize the deterioration of the economy.
b. Responding to the political interference in medicine pricing decisions.
c. Fear of a negative reaction from citizens if the prices of medicines increase.
d. The political affiliation of the general secretaries of the NMPB makes them try to please politicians and to satisfy the media by hiding the real prices of medicines, and thereby creating a good image and positive public opinion.
e. The rule of Gargoush^1^, which is an expression of injustice and the rule of the strong over the weak.

## Discussion

Throughout the world, medicines are one of the key elements in the successful modern health care system^(10)^. The cost of medicines is a huge burden on health expenditure in all countries. In Sudan, NMPB has adopted a number of measures to control continuous skyrocketing medicine prices that resulted from devaluation of the national currency. Pre- and post-marketing quality control laboratory tests are important to ensure the quality of medicines and to detect SF products throughout the supply chain. The WHO has stressed the need for pre-marketing quality testing^(6)^. Until 2010 and in line with WHO guidelines^(6)^ and its bylaws of medicine registration 2009, NMPB approval for a medicine required, among others, the passing of laboratory pre-test for quality. Although these samples might or might not have been produced by the manufacturer that provided them, the report of the NMPB showed that 587 (17%) samples failed the quality testing in 2009. The challenge for Sudan is that India, Pakistan, Egypt and China are among the top sources of registered medicines in Sudan with 1,040 out of a total of 3,873 registered items. These countries are sources of the majority of reported cases of falsified medicines that have been seized^(11–13)^.

To maximize the benefit from the limited resources available for the NMPB, there should be an added value from the registration of more alternatives of the same generic medicine. The literature reveals that there would be no monopoly if more than 4 competitors are on the market^(14)^. Additionally, in pharmaceutical sector, the competition was most effective with institutional purchasers, who are price conscious and have enough expert capacity to procure medicines, but is less effective if individuals are the main purchasers. This is because patients cannot safeguard their interests for a number of reasons that include, but not limited to, asymmetry of information, price inelasticity of demand and trust^(15)^.

Despite increased number of registered generic medicines, the problem of access to potentially affordable medicines was not solved. This is mainly because most of newly registered medicines are already on the market from different sources. Secondly, the root causes of the high prices of medicines were not the lack of competition; rather the scarcity of foreign currencies and the new NMPB’s pricing policies.

It is clear that the lack of strategic vision, delays in government decisions to keep pace with the official exchange rate, and failure to avail foreign currencies for medicine importation have led to stagnation in the pharmaceutical market in Sudan. The majority of pharmaceutical companies have ceased to operate and some foreign companies have been forced to quit the market. However, some companies have continued to operate, despite the large difference between the exchange rate set by the NMPB and the official exchange rates for foreign currencies, which companies could have not been able to obtain to import medicines. Some importers and in order to meet their ethical obligations and achieve the sales’ target set by their foreign manufacturers, secure their foreign currencies from the black market at higher exchange rates.

The findings of this study are in line with the literature, which showed that adoption of stringent unrealistic measures to reduce price of medicine, regardless of losses caused to the pharmaceutical industry, leads to sever medicine shortages, increased the prices unofficially, creates a black market and risks the patients’ life^(16, 17)^. In Sudan, the insistence of the government to use an exchange rate, which is not existent, has had a negative impact on pharmaceutical availability. For instance, the value of the imported medicines in the first six months of 2019, according to the Central Bank of Sudan, dropped from US$90 million to only US$10 million in the same period in 2020. The situation in locally produced medicines is, by no means, not better. The Federal Ministry of Health (FMOH) reported that the quantities that produced in 2019 were only 8% of those produced in 2018. The consequences are low medicine availability, increased number of illegal traders, and patients secure their health needs from unofficial sources of medicines at higher prices. This trend will endanger patients’ lives.

This study discussed the impact of the updating of the requirements for the registration of medicines and the pricing system of the registered pharmaceuticals in Sudan during the past 10 years from the perspective of the members of the SSRAP. The paper recommends that the medicines registration requirements need to be improved based on the WHO guidelines (for example^(6, 18)^). Further, to reduce the registration time and to prioritize the registration of generic medicines, NMPB could rely on stringent regulatory authorities on the WHO list, such as European Medicines Agency, Food and Drug Administration in the United States of America in the United State of America, and the WHO prequalification program. Literature shows that reliance on well-established stringent regulatory authorities reduces the resources needed for medicines registration and shortens the process from years to months in the Caribbean region^(19)^. Moreover, the adoption of the CTD format will contribute to the robust accelerated registration of safe, effective and quality products. However, the application of the CTD will require a huge investment in the training of NMPB technical staff on how to critically appraise the CTD. Lastly, the reliance on the laboratory quality testing alone does not guarantee the quality of medicines with regard to stability and bioavailability, since pharmacopoeial specifications do not necessarily address these issues^(20)^.

The efforts of the successive governments to allocate foreign currencies for medicines at the official exchange rate have failed. However, the NMPB’s intransigence has often led to the imposition of a lower exchange rate than the official one in order to reduce the cost of medicines in order to alleviate the suffering of patients. This noble goal has practically been impossible and turned out into a disaster that threatens the health system in Sudan because it leads to a chronic shortage of medicines. Reference to this empirical experience that lasts for more than 8 years, the authors recommend liberalization of the exchange rate of foreign currencies, which does not mean liberalization of medicine prices. It will enable medicine companies to access the foreign currencies needed to supply medicines without making losses.

A fair pricing bylaws needs to be developed and approved, because medicine affordability is a critical factor in a country like Sudan, where over 45% of the population lives below the poverty line and, according to the FMOH, 70% pay for their health services from their own pockets. The fair pricing system will encourage the growth of the pharmaceutical industry without compromising patients’ access to medicines^(1)^. The medicine affordability cannot be achieved by obliging pharmaceutical companies to sell their medicines at an exchange rate below or equal to the official rate, while the foreign currencies at these rates are not existent.

### Limitations

The paper must be understood within the limitations of being written by the president of the SSRAP and ex-members of staff of the NMPB. Secondly, to compensate the inbuilt bias resulting from the participants who were members of the SSRAP, the majority of the questions were designed in the format of multiple choices.

## Conclusion

The study highlighted the importance of considering options for balancing patient access to quality medicines, and reasonable pricing policies that encourage the national pharmaceutical industry to flourish and maintain a steady flow of quality-assured medicines from abroad to the local market. Also reliance on stringent regulatory authorities on the WHO list, and adoption of the CTD format were reported to be an efficient way of improving medicines registration. However, reliance on laboratory quality testing alone was not seen to be effective in safeguarding people against FS medicines.

## Declarations

### Ethics approval

As the study is descriptive by nature, with no diagnostic or clinical intervention, it was determined to be exempted from review by the institutional review board at the Federal Ministry of Health, Sudan. However, the General Directorate of Research, Federal Ministry of Health, had reviewed and approved the study protocol.

### Consent to participate

Verbal informed consent was obtained from the participants for their anonymized information to be published in this article.

## Data Availability

All relevant data are within the manuscript and its Supporting Information files

## Footnotes

1 The Story tells Mr. Gargoush was an unjust ruler. Once he visited the place where the criminals were executed. The jailer was confused about the execution by hanging in one of the convicts because his neck was short and larger than the rope tie. The unjust ruler Mr Gargoush, chose one of those who came to see the execution of the sentences, and ordered the jailer to execute him instead of the criminal.

